# TRAF1 S146 is constitutively phosphorylated in primary CLL cells by PKN1/2

**DOI:** 10.64898/2026.02.11.26346036

**Authors:** Birinder Ghumman, Laura Nicolucci, Tania H. Watts, Ali Abdul Sater

## Abstract

TRAF1 is a pro-survival signaling adaptor that contributes to NF-κB activation downstream of a subset of TNFR superfamily members. TRAF1 is overexpressed in many cancers of mature B cells, including chronic lymphocytic leukemia (CLL). Previous studies have established that TRAF1 S146 is a target of phosphorylation by the kinase PKN1 and that PKN1 is required to prevent cellular inhibitor of apoptosis protein (cIAP)-dependent degradation of TRAF1 in the CD40 signaling complex. The kinase inhibitor OSST167 inhibits PKN1 in the nm range and its addition to primary CLL cells was shown to induce dose-dependent loss of TRAF1 and concomitant increases in activated caspase 3 and cell death. These studies identified PKN1 as a target for therapy of CLL. To identify more potent and specific PKN1 inhibitors for therapy of B cell cancers it is important to measure a direct target of PKN1, such as phospho-TRAF1. To this end, here we use overexpression of an S146A mutant of human TRAF1 in 293 cells to validate a recently generated phospho-TRAF1 S146-specific antibody and to confirm that this phosphorylation is lost upon treatment with OTSSP167. Using Cas/Crispr knockout in RAJI cells we also show that both PKN1 and the closely related family member PKN2 can phosphorylate TRAF1 S146. We further show that TRAF1 S146 is constitutively phosphorylated in primary human CLL cells, including those with p53 mutations and that this phosphorylation is sensitive to inhibition with OTSSP167. These findings provide support the development of more potent PKN1/2 inhibitors for CLL.

## Introduction

Chronic lymphocytic leukemia (CLL) is the most prevalent adult leukemia. In recent years, first line CLL therapy has dramatically shifted towards targeted therapies including drugs such as Ibrutinib to inhibit BCR signaling or venetoclax to target cellular survival through antagonizing Bcl-2 (for review see ^1^). Despite the promise of these new therapies, resistance still occurs and there is a need for additional approaches to target venetoclax resistant CLL survival signaling.^2^

TRAF1 is an NF-κB inducible signaling adaptor that contributes to feedback enhancement of NF-κB signaling downstream of several TNFR superfamily members.^3^ TRAF1 is generally undetectable in resting normal immune cells but overexpressed in cancers of mature B cell origin, including CLL. TRAF1 has been directly implicated in B lymphomagenesis using mice expressing constitutively active NF-κB2, where deletion of TRAF1 restored normal B cell homeostasis. TRAF1 forms a complex with TRAF2 and the cellular inhibitors of apoptosis proteins (cIAPs) ^5^ to increase NF-κB activation downstream of TNFRs such as TNFR2, CD40, CD30, and 4-1BB.^6-9^ This in turn leads to induction of Bcl-2 family members such as Mcl-1 that can contribute to venetoclax resistance. ^2,3,10^

TRAF1 Serine 146 (S146) is a target of phosphorylation by the protein kinase C related kinase PKN1.^11^ We previously showed that in the absence PKN1, TRAF1 is targeted for degradation by cIAPs during CD40 signaling.^10^ Conversely, phosphorylation by PKN1 allows TRAF1 protein levels to be maintained during TNFR signaling, resulting in increased NF-kB signaling and induction of prosurvival Bcl-2 family members.^10^ Previous work identified the kinase inhibitor OTSSP167 as inhibiting PKN1 with an IC_50_ of 18nm.^10^ When added to primary CLL cells, OTSSP167 led to a dose-dependent loss of TRAF1 protein, with concomitant loss of Bcl-2 and Mcl-1, and increased levels of activated caspase 3, leading to cell death.^10^ Moreover, OTSST167 synergized with venetoclax in inducing CLL cell death.^10^ These findings identified PKN1 as a potential target for inhibition for CLL. However, the OTSSP167 inhibits other kinases, notably MELK ^12^ with an IC_50_ in the picomolar range. Thus, it is of interest to find more selective PKN1 inhibitors. As TRAF1 is regulated both transcriptionally through NF-κB and at the level of protein stability through PKN1, it would be preferable to measure phospho-TRAF1 directly rather than total TRAF1 to screen for improved PKN1 inhibitors. To this end, in this study, we used 293 cells over-expressing WT or S146A TRAF1 to validate a recently generated antibody specific for the phospho-S146 form of TRAF1.

PKN1 is part of a family that includes 3 related kinases PKN1, 2 and 3, with PKN1 and 2 reported to be ubiquitously expressed and 50% identical at the amino acid level.^13^ Here we show that either PKN1 or PKN2 can phosphorylate TRAF1 on S146. We also show that TRAF1 S146 is constitutively phosphorylated in primary patient CLL cells and that this phosphorylation is inhibited by OTSSP167, previously identified as inhibiting PKN1.^10^ Together our studies, suggest constitutive PKN1/2 activity in CLL cells and provide a means of monitoring this activity through a phospho-TRAF1 S146-specific antibody.

## Materials and Methods

### Human subjects

Peripheral blood mononuclear cells from patients with a diagnosis of CLL were obtained from the Leukemia Tissue Bank at Princess Margaret Cancer Centre/University Health Network (UHN), Toronto, Ontario. Informed consent for tissue bank donation was in compliance with the Declaration of Helsinki and in agreement with the UHN Research Ethics Review Board (protocol # 01-0573). Research with these samples was conducted at the University of Toronto, with approval of the University of Toronto ethics board (protocol # 60791).

### Cell lines

Human embryonic kidney 293 cells (ATCC) were cultured in Eagle’s Minimum Essential Medium with Earle’s Salts (purchased from Wisent Inc) supplemented with 10% Fetal Calf Serum (FCS, Wisent Inc) and 1% of 100X Glutamine-Penicillin-Streptomycin (GPS) (Sigma-Aldrich, Oakville, Canada). RAJI cells (ATCC) were cultured in RPMI with 25mM Hepes (purchased from Wisent Inc.) supplemented with 10%FCS from Wisent and 1% of 100X Glutamine-Pyruvate Penicillin-Streptomycin (GPPS) (Sigma-Aldrich, Oakville, Canada). OP-9 stromal cells were kindly provided by J.C. Zuniga-Pflücker, Sunnybrook research institute, Toronto, Canada. Cell lines tested negative for mycoplasma (Mycoplasma detection kit, Millipore-Sigma, Oakville, Ontario).

### Antibodies

Anti-TRAF1 S146 [EPR25987-11] - BSA and Azide free was kindly provided by Abcam, Cambridge, UK. Anti-TRAF1 (Cell Signaling Technology (CST) clone 45D3, cat # 4715), anti-PKN2 (CST cat#2612) and anti-GAPDH (CST cat# 2118) were purchased from New England Biolabs (NEB), Whitby, ON. Anti-PKN1 (cat# 610687) was purchased from BD Biosciences, until it was discontinued, after which we used anti-PKN1 from Thermo Fisher Scientific (Cat# PA587454) purchased from Life Technologies Canada, Burlington, ON. Goat anti-Rabbit-HRP and Goat anti-mouse-HRP from Jackson Immunoresearch were purchased from Cedarlane. Goat anti-Actin-HRP (A3854) was purchased from Millipore-Sigma Canada.

### Knockout of PKN1 and PKN2

Cell lines with gene disruptions in PKN1 and/or PKN2 were generated at the Genomic Engineering and Molecular Biology (GEMb) core facility in the Faculty of Medicine at the University of Ottawa (RRID: SCR_022954). Briefly, guide sequences targeting gene ID 5585 (PKN1) or 5586 (PKN2) were cloned into pLentiCRISPRv2 (puromycin resistant) and pLentiCRISPRv2BLAST (blasticidin resistant), respectively. A non-mammalian targeting guide sequence targeting Renilla luciferase was cloned into both these vectors as a control. Guide sequences were chosen to minimize the potential of off-target cutting^14^ and cloned as previously described.^15^ Guide sequences are listed in Table 1. Lentiviral particles were generated as previously described^15^ and were used to transduce RAJI cells, followed by selection with puromycin (1 μg/ml) or blasticidin (5 μg/ml) as appropriate. Editing efficiencies for guides targeting gene ID 5585 or 5586 were measured by generating PCR products flanking the edit sites using primers 5’-CTTAATGTGGGGGACGCTGT & 5’-TCTTGAGTGTTACCGGGTGC or 5’-AACTGCTGATCCAGACGTGTT & 5’-AATCGCCTTAGCTGTTAGCG, respectively, and sequencing these primers using Oxford Nanopore technology (sequencing performed by Plasmidsaurus). Subsequently editing efficiencies were measured using the CRISPRESSO2 softwaresuite,^16^ subtracting sequencing noise measured in the control population. Independent pairs of guides were then used to generate lines in which both genes were disrupted (5585 g2+5586 g1 & 5585 g3+5586 g3). Editing rate represents a snapshot of the polyclonal population at one point in time and likely increased over passaging from this initial measured value as the lines stably express the CRISPR/Cas9 system.

**Table I:**
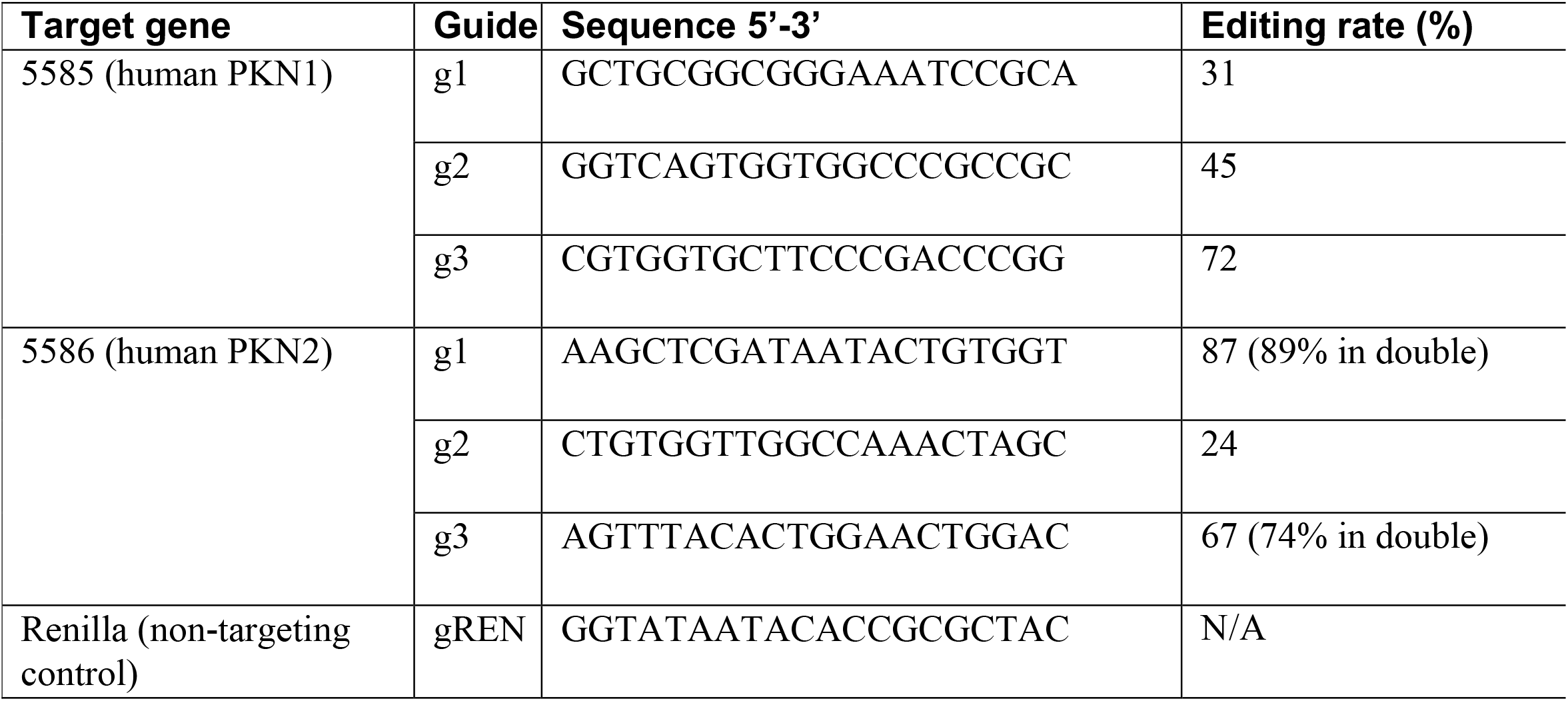
Editing rates for PKN1 and PKN2 Guide sequences and Nanopore-measured editing rates. Where editing rates in double mutants are expressed, this is measuring efficiency of the 5586 editing system introduced into the lines already containing the 5585 editing system.

### Western Blot

Media was removed and cells were washed once with cold PBS (Wisent Inc), then lysed for protein extraction. Cells were lysed in RIPA lysis buffer (50mM Tris-HCl pH 8.0, 150mM NaCl 1.0% Nonidet P-40, 0.5% Deoxycholate, 0.1%SDS (Millipore-Sigma, Oakville, Canada) with phosphatase and protease inhibitor mix (Roche, Basel, Switzerland). Total protein concentration was quantified by a colorimetric assay (Bio-Rad, Berkeley, United States), then subjected to SDS-PAGE (4-20% gradient gel) and transferred to polyvinylidene difluoride membranes (wet transfer; transfer with 10% methanol, Bio-rad). After blocking with 5% NFDM in TBST (TBS with 0.1%Tween), membranes were probed with antibodies specific for pS146 of TRAF1 (Abcam clone EPR25987-11), TRAF1 (clone 45D3, Cell Signaling, Danvers, United States) for 1hr at rm temp, PKN1 (BD Biosciences, Franklin Lakes New Jersey, United States) overnight at 4C and GAPDH for 30 min at rm temp, followed by HRP-conjugated anti-rabbit or anti-mouse antibody (Jackson Immunoresearch, Baltimore, United States), and signals were detected with a chemiluminescence substrate (Clarity Western ECL substrate from Bio Rad Canada and Immobilon Forte Western HRP substrate from Millipore Sigma Canada).

### Primary CLL cell culture and treatment with inhibitors

OP9 cells were re-suspended at 10^5^ cells/mL in OP9 Media (α-MEM from Thermo Fisher Scientific with added 20% FCS, glutamine, penicillin and streptomycin) and seeded at 5×10^4^ cells per well into a 24-well plate. When 80-90% confluent CLL patient samples (PMBC) were thawed and re-suspended at 8×10^6^ cells/mL in high glucose complete media (RPMI 1640 supplemented with 2.5g/L total glucose and other additions as indicated for RAJI cells above). The OP9 media was removed and CLL cells were plated at 2×10^6^ cells per well on a 24-well plate containing confluent OP9 and rested overnight. Samples were treated with OTSSP167 (dose indicated in the figures) or 0.1% DMSO media control for 24hrs, then samples were gently removed from the well and prepared for Western blot as described above.

### Data availability

Raw data will be made available upon request.

## Results

### Validation of pTRAF1 specific antibody Abcam

Anti-phospho-TRAF1 S146 (clone EPR25987-11 Abcam) is reported to bind phospho-TRAF1 containing peptide and detects TRAF1 protein in RAJI cells. To validate that this antibody is specific for the phospho-S146 form of TRAF1 and not other potential sites on TRAF1, we used parental 293 cells which do not express TRAF1, as well as 293 cells transfected with WT or a mutant form of human TRAF1 in which serine 146 is replaced with alanine (S146A TRAF1) (Figure 1A). The results show that anti-phospho-TRAF1 antibody detects WT but not S146A TRAF1. We next used OTSSP167, a kinase inhibitor previously shown to inhibit PKN1,^10^ to determine if total TRAF1 as well as phospho-TRAF1 S146 was sensitive to this inhibitor (Figure 1 B-D). We found that administration of OTSSP167 resulted in loss of pTRAF1 S146 with an IC50 of 29.4nm (Figure 1 B, D, E).

**Figure 1:**
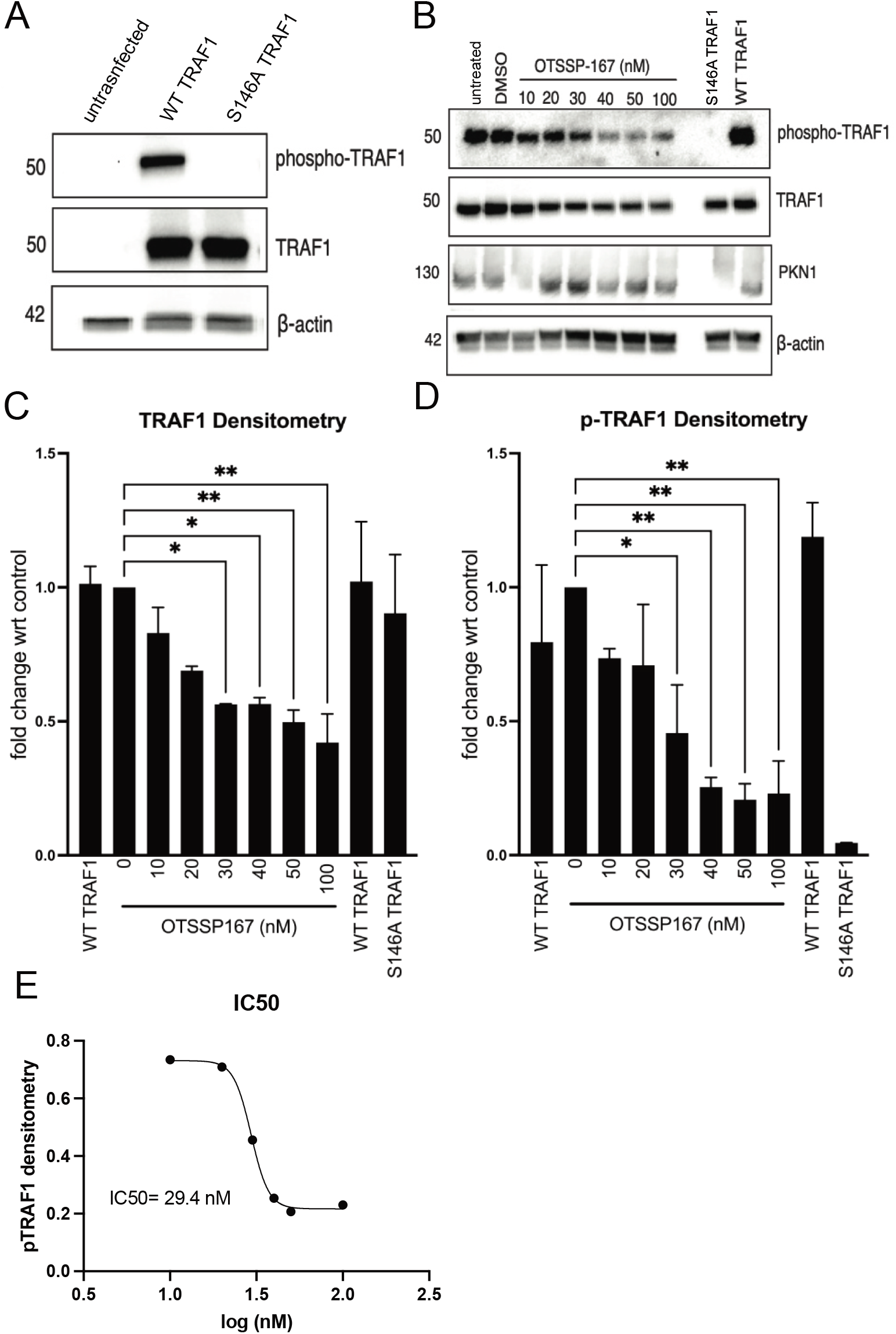
OTSSP167 inhibits TRAF1 phosphorylation in a dose dependent manner. (**A**) 293 cells were either untransfected or transfected with wildtype TRAF1 (TRAF1 WT) or S146A mutant TRAF1 (TRAF1 S146A). Whole cell lysates were then immunoblotted for p-S146-TRAF1 (p-TRAF1), total TRAF1 (TRAF1), and β-actin (data are representative of at least 3 independent experiments). (**B**) 293 cells overexpressing WT TRAF1 were treated with 10-100 nM OTSSP167 for 24 h. Whole cell lysates were then immunoblotted for p-S146-TRAF1 (p-TRAF1), total TRAF1 (TRAF1), PKN1, and β-actin. Lysates from TRAF1 WT and S146A overexpressing 293 cells were also included on the blots as controls for p-TRAF1 assay. (**C-D**) Densitometry of TRAF1 and p-TRAF1 band intensity from 3 independent experiments, as in panel B, were quantified by ImageLab software and (**E)** used to calculate the IC_50_ for PKN1 based on reduction of expression of pTRAF1 S146.

### PKN1 and PKN2 both contribute to TRAF1 phosphorylation at S146

PKN1 and PKN2 are highly similar and both are expressed by RAJI lymphomas (**Figure 2**). Therefore, to determine whether PKN2 can also contribute to phosphorylation of TRAF1, we generated mutant versions of RAJI in which PKN1 or PKN2 or both were knocked out by Cas/Crispr technology. Knockout of either PKN1 or PKN2 each partially reduced the level of pTRAF1, with additive effects of knockout of both kinases (Figure 2). These data suggest that the highly similar PKN1 and PKN2 can each contribute to TRAF1 S146 phosphorylation.

**Figure 2:**
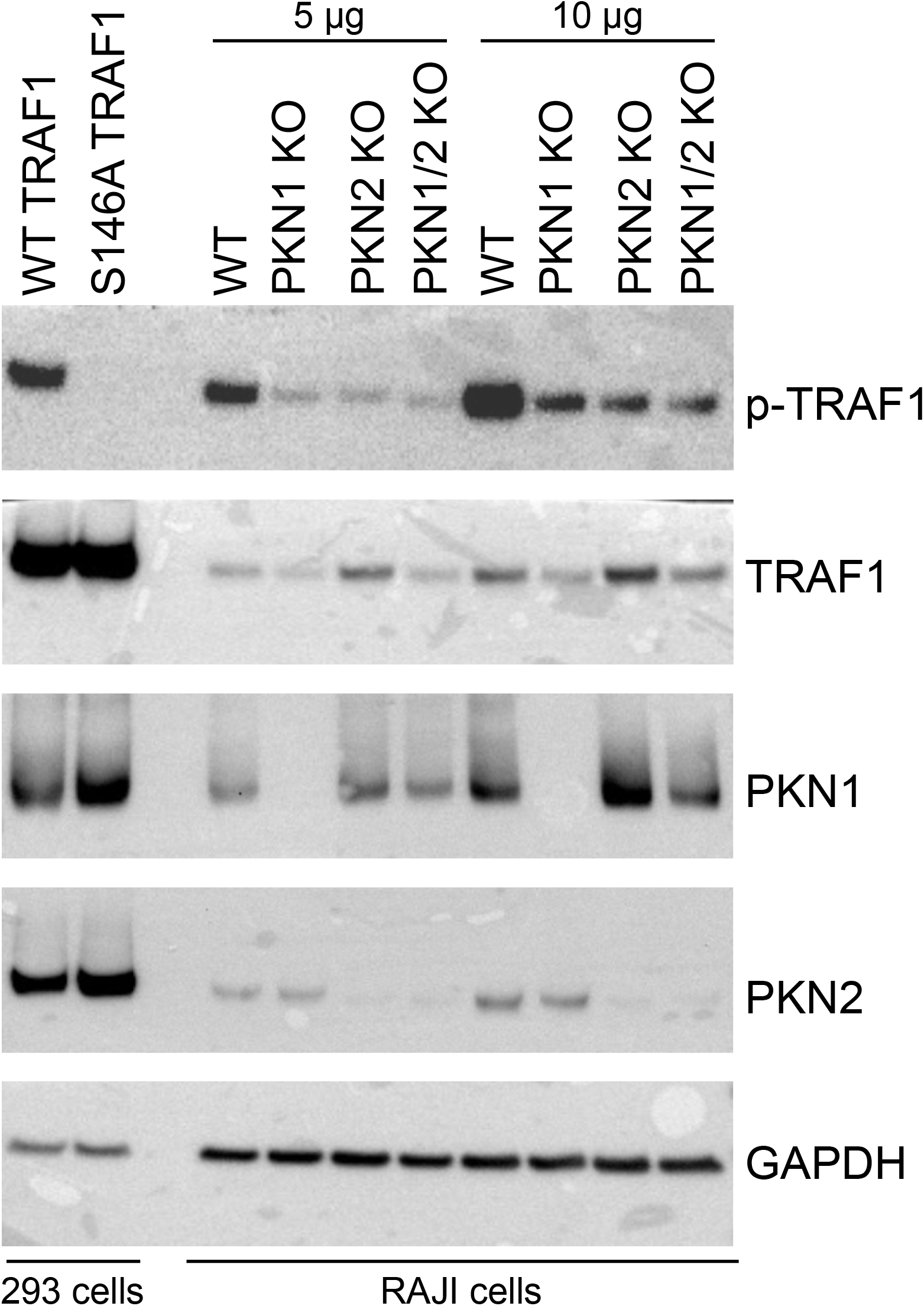
PKN1 and PKN2 are required for TRAF1 phosphorylation. 5 or 10 µg of whole cell lysates from WT, bulk PKN1 knockout (PKN1 KO), bulk PKN2 (KO) or bulk PKN1 and PKN2 double knockout (PKN1/2 KO) RAJI cells were immunoblotted for pTRAF1 S146 (p-TRAF1), total TRAF1 (TRAF1), PKN1, PKN2 and GAPDH. Lysates from TRAF1 WT and S146A overexpressing 293 cells were loaded separately as controls for pTRAF1 assay. These data are representative of 3 similar experiments, in which 2 of 3 experiments showed similar role for PKN1 and PKN2 and one experiment showed greater role for PKN1 than PKN2 in TRAF1 S146 phosphorylation.

### TRAF1 S146 is constitutively phosphorylated in primary CLL cell lines

CLL cells have been reported to have constitutive signaling through TNFRs such as CD40 and CD30, resulting in constitutive NF-κB activation,^17,18^ which induces TRAF1^7^ as well as Bcl-2 and Mcl-1.^10^ To determine whether TRAF1 is phosphorylated in CLL cells, we conducted western blot analysis of PBMC from CLL patients, after overnight culture of the cells with OP9 stromal cells. The majority of cells in the PBMC are CLL cells and our previous results have shown that neither OP9 stromal cells or resting normal lymphocytes express TRAF1.^10^ We analyzed CLL cells expressing p53, which has a poor prognosis, CLL cells with the 13q translocation, which has a more favourable prognosis, as well as CLL samples of undetermined genotype (**Figure 3A**). The results show that phospho-TRAF1 is constitutively present in all CLL patient samples analyzed, albeit with some variably between samples. Moreover, this phosphoTRAF1 signal was sensitive to treatment with OTSSP167, with an IC_50_ of 39 nM (**Figure 3 B-F**).

**Figure 3:**
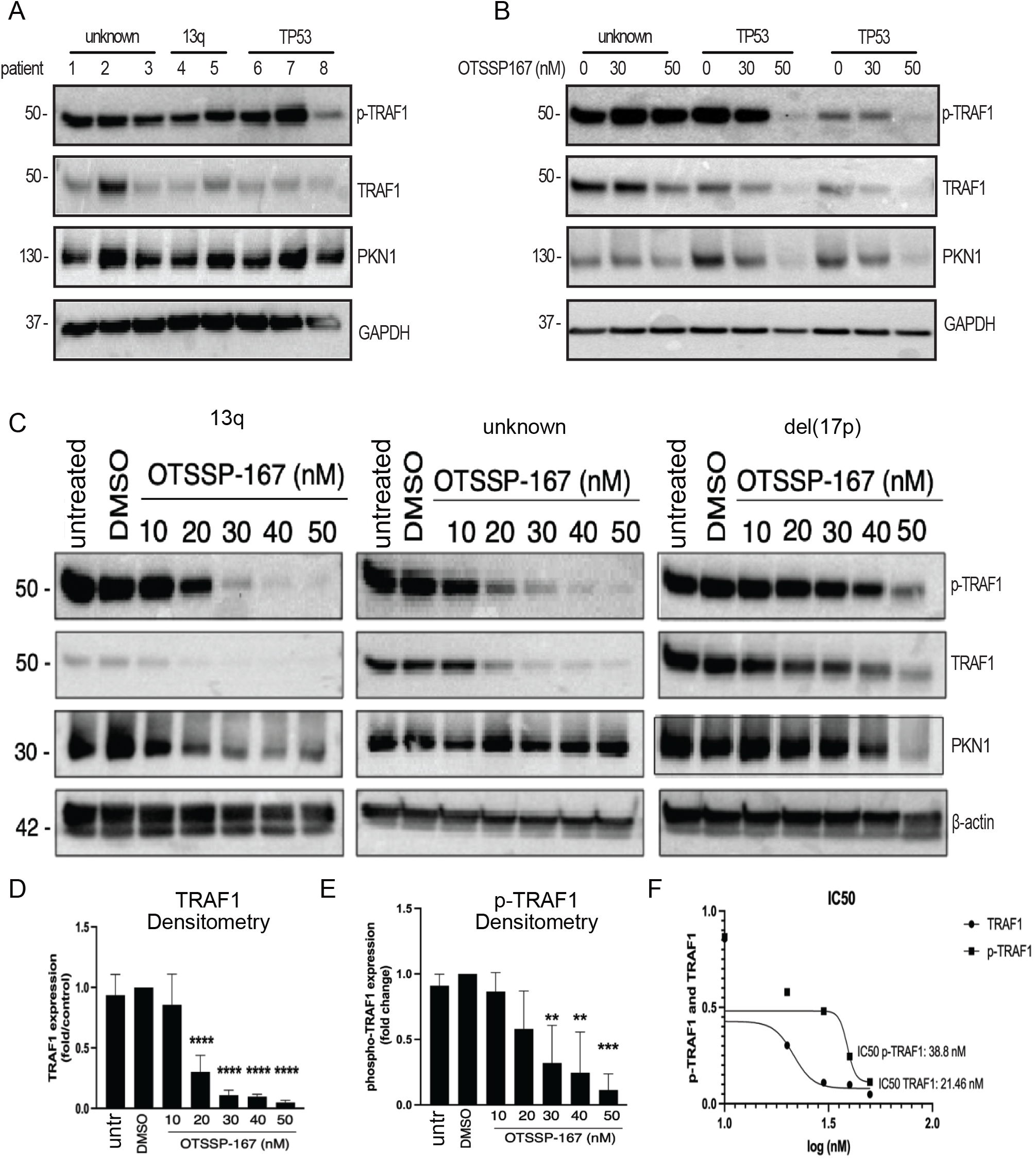
OTSSP167 decreases total and phospho-specific TRAF1 protein levels in primary CLL cells. (**A**) Lysates from CLL cells from patients with unknown (n=3 patients), 13q (n=2 patients) or p53 mutations (TP53) (n=3 patients) were subjected to western blot analysis for phospho-S146 specific TRAF1 (p-TRAF1), total TRAF1 (TRAF1), PKN1, or GAPDH. (**B**) CLL cells from representative donor from an unknown or two different TP53 groups from panel A were treated with DMSO control or OTSSP167 for 24h at the indicated concentrations. Lysates were then immunoblotted for phospho-TRAF1, total TRAF1 (TRAF1), PKN1 or GAPDH. (**C**) CLL patient cells with unknown, 13q mutations, or 17p deletion (del(17p); results in loss of WT p53) were treated with OTSSP167 at doses ranging from 10-50 nM for 24h. Lysates were then immunoblotted for phospho-TRAF1, total TRAF1 (TRAF1), PKN1 or β-actin. (**D-E**) Densitometry of TRAF1 (**D**) and p-TRAF1 (**E**) band intensity from 3 CLL patient cells, as in panel C were quantified by ImageLab software. (**F**) IC_50_ of total TRAF1 (TRAF1) and phospho-specific TRAF1 (p-TRAF1) was calculated from densitometry analysis shown in panels D and E.

## Discussion

The results presented in this study validate that a phospho-S146-specific antiTRAF1 antibody raised against a phospho-peptide (Abcam clone EEPR25987-11) is specific for the phosphate modification of S146 on the intact TRAF1 protein in cells. Using gene knockout in the RAJI lymphoma cell line we show that both PKN1 and PKN2 can contribute to this phosphorylation. The kinase inhibitor OTSSP167 has a pM affinity for MELK,^12^ but an IC_50_ for PKN1 in the nM range.^10^ In cells, we previously showed that the IC50 for TRAF1 loss upon inhibition with OTSSP167 was around 30 nM, and similar to the IC_50_ for PKN1.^10^ Here we show that treatment of 293 cells or primary CLL with OTSSP167 results in loss of the S146 phosphate group on TRAF1, with a similar IC_50_. These data are consistent with PKN1 phosphorylating TRAF1 at serine 146 in primary CLL to promote TRAF1 stability. Our previous study showed that loss of TRAF1 in response to OTSSP167 resulted in loss of Bcl2, Mcl-1, increased activated caspase 3 and cell death, with a similar IC_50_ for each readout.^10^ As PKN1 and PKN2 are 50% identical in their NTD binding domain, the location of OTSSP167 binding, it seems likely that OTSS167 is targeting both kinases. Taken together, these studies validate phospho-TRAF1 staining as a readout for inhibition of PKN1/2 in cell lines as well as primary CLL cells. This antibody should therefore be useful in screening new PKN1/2 inhibitors for on target effects in CLL.

Our previous work used loss of total TRAF1 protein in response to OTSSP167 to show that PKN1 is important for TRAF1 stability during constitutive signaling in primary CLL cells. Here we validate this through direct measurement of phospho-TRAF1. We also provide evidence here that TRAF1 is constitutively phosphorylated in primary CLL independent of CLL prognosis.

## Data Availability

All data produced in the present study are available upon reasonable request to the authors

## Acknowledgements

We thank Andrea Arruda and Mark Minden and the PMH tumor bank for access to samples and Abcam for providing the phospho-TRAF1-specific antibody prior to commercial release. We thank the Genomic Engineering and Molecular Biology (GEMb) core facility in the Faculty of Medicine at the University of Ottawa (RRID: SCR_022954) for generation of PKN1 and PKN2 knockout cells. plentiCRISPR v2 was a gift to the GEMb facility from Feng Zhang (Addgene plasmid # 52961; http://n2t.net/addgene:52961; RRID:Addgene_52961). plentiCRISPR v2-Blast was a gift from Mohan Babu (Addgene plasmid # 83480; http://n2t.net/addgene:83480; RRID:Addgene_83480).

## Author Contributions

THW and AAS conceptualized and obtained funding for the study and wrote and edited the manuscript. B.G. conducted the investigations and edited the manuscript. LN assisted with investigation and manuscript preparation.

